# Development and Feasibility of a Primary Care Provider Training Intervention to Improve Atrial Fibrillation Management

**DOI:** 10.1101/2023.03.21.23287553

**Authors:** J. Michael Daw, Tiffany Armbruster, Zack Deyo, Jennifer Walker, Lindsey A Rosman, Samuel F. Sears, Anthony J. Mazzella, Anil K. Gehi

## Abstract

**Background:** Disparities in atrial fibrillation (AF) care are partially attributed to inadequate access to providers with specialized training in AF. Primary care providers (PCPs) are often the sole providers of AF care in under-resourced regions.

**Objective:** To create a virtual education intervention for PCPs and evaluate its impact on use of stroke risk reduction strategies in AF patients.

**Methods:** A multi-disciplinary team mentored PCPs on AF management over 6 months using a virtual case-based training format. Surveys of participant knowledge and confidence in AF care were compared pre- and post-intervention. Hierarchical logistic regression modeling was used to evaluate change in stroke risk reduction therapies among patients seen by participants before or after training.

**Results:** Of 41 participants trained, 49% worked in family medicine, 41% internal medicine, and 10% general cardiology. Participants attended a mean of 14 one-hour sessions. Overall, appropriate use of oral anticoagulation (OAC) therapy (CHA_2_DS_2_-VASc score ≥1 men, ≥2 women) increased from 37% to 46% (p<.001) comparing patients seen pre- (n=1739) to post- (n=610) intervention. Factors independently associated with appropriate OAC use included participant training (OR 1.4, p=.002) and participant competence in AF management (by survey). Factors associated with decreased OAC use included patient age (OR 0.8 per 10 years, p=.008), nonwhite race (OR 0.7, p=.028). Provider knowledge and confidence in AF care both improved (p<.001).

**Conclusions:** A virtual case-based PCP training intervention improved use of stroke risk reduction therapy in outpatients with AF. This widely scalable intervention could improve AF care in under-resourced communities.

**CONDENSED ABSTRACT:** A virtual educational model was developed for primary care providers to improve competency in AF care in their community. Following a 6-month training intervention, the rate of appropriate oral anticoagulation (OAC) therapy among patients cared for by participating providers increased from 37% to 46% (p<.001). Among participants, knowledge and confidence in AF care improved. These findings suggest a virtual AF training intervention can improve PCP competency in AF care. This widely scalable intervention could help improve AF care in under-resourced communities.

## INTRODUCTION

Atrial fibrillation (AF) care is complex and often sub-optimal. Prior studies have demonstrated a lack of adherence to AF guidelines, including low rates of anticoagulation, disparate use of rhythm control medications, and high rates of hospitalizations.^1-3^ The prevention of stroke is a mainstay of AF management, yet prior studies have suggested a significant knowledge gap among providers regarding therapies for stroke risk reduction with AF.^1,4,5^ Moreover, rural and under-resourced regions are disproportionately affected by sub-optimal care due to resource limitations and inadequate access to community providers with specialized training in AF, resulting in greater non-adherence to clinical guidelines, underutilization of stroke prophylaxis, and higher rates of in-hospital mortality compared to urban communities.^6–10^

Primary care providers are often the sole providers serving these vulnerable patient populations and may have difficulty gaining confidence in AF care as management recommendations become increasingly complex. Disparities of access to specialty AF care may be more adequately addressed through engaging with primary care providers who can become local AF care partners. Numerous studies show that “hub-and-spoke” networks using the Project ECHO model (Extension for Community Healthcare Outcomes) can effectively reduce health disparities in under-resourced regions.^15–22^ Whether this model of care can be implemented to improve access to high quality AF care is unknown.

Accordingly, we sought to develop a group-based virtual education intervention for primary care providers using the Project ECHO model and to evaluate its early impact on the prescription of oral anticoagulation and antiplatelet therapy for stroke risk reduction, as well as overall provider competency in AF care.

## METHODS

### Provider Population

Providers within the University of North Carolina Health system were recruited through direct requests for participation and general requests to the community through flyers at local CME programs and email. Enrollment in the intervention was open to healthcare providers with any degree (physician, nurse practitioner, or physician assistant) practicing family medicine, internal medicine, or general cardiology who cared for at least >10 patients with AF in a year in the primary care or general cardiology setting. All provider participants gave written informed consent for inclusion in the study and were trained on the curriculum agenda, participant expectations, evaluations, and protocols regarding patient confidentiality during case presentations. The study received approval by the Institutional Review Board at the University of North Carolina at Chapel Hill.

### Design of the Intervention

We based our intervention on the Project ECHO model developed at the University of New Mexico to reduce health disparities in under-resourced and remote regions. The Project ECHO model creates a “hub-and-spoke” knowledge-sharing network between specialists and regional care providers (**Central Illustration**). The ECHO model aims to maximize the capacity of front-line providers by using technology to leverage scarce resources in under-resourced areas. Through virtual group-based mentoring, known as the “Tele-ECHO Clinic,” participants learn and receive feedback on implementation of “best practice” recommendations of AF care through case-based learning.

We developed a curriculum for participants including comprehensive AF and comorbidity management as well as EKG interpretation skills. The curriculum involved weekly Tele-ECHO training sessions led by a trained electrophysiologist (AKG) and supported by a multi-disciplinary team of content experts including cardiology, sleep medicine, endocrinology, pharmacy and clinical psychology providers. The curriculum predominantly utilized a case-based format supplemented with didactics and training in EKG interpretation. The intervention was held via real-time interactive videoconferencing over two six-month periods from May 2019 to October 2019 and January 2020 to June 2020. Notably, the flexible format of the intervention allowed uninterrupted delivery during the early phase of the COVID-19 pandemic. Participants were asked to attend at least 12 of the 24 available sessions (receiving CME credit for each hour of participation) and to present a minimum of two deidentified AF case studies from their own practice.

Each Tele-ECHO training session began with introductions of all participants followed by a review of a unique or interesting EKG. Subsequently, an AF case was presented by a participant and management options were discussed with group participation guided by the facilitator. Recommendations were summarized verbally at the conclusion of the case presentation, transcribed, and forwarded in writing to the participant whose case was discussed. The Tele-ECHO training session concluded with a 20-minute formal didactic lecture presented by one of the content experts (Supplement).

### Patient Population

The effect of our intervention on AF provider management was assessed by evaluating the prescription of stroke risk reduction therapy in AF patients managed by providers participating in the intervention. Patients included in the cohort included those with a diagnosis of AF who were seen by one of the participants within one year before or one year after the intervention. Only one outpatient encounter per patient was included in the analysis and patients were categorized as being seen in either the pre- or post-intervention period. Any patient with AF seen during the 6-month intervention was excluded from analysis. If patients were seen both during the pre- and post-intervention periods, only the first (pre-intervention) encounter was included in analysis.

### Patient Characteristics

Patient demographics, medical history, and medications were abstracted from the Carolina Data Warehouse for Health (CDW-H), which stores electronic health record databases for all University of North Carolina (UNC) facilities and affiliated hospitals. The CDW-H uses a standardized HIPAA-compliant data dictionary which is harmonized with discrete data from electronic health records through validated automated computer algorithms and standard methodology.^25^ Harmonized databases include patient demographics, insurance status, and clinical information from all inpatient and outpatient visits to UNC and affiliated facilities.

Sociodemographic information was collected closest to the training intervention. The social deprivation index (SDI), which calculates a composite score of socioeconomic factors associated with poor access to healthcare and health outcomes based on patient ZIP code, was assessed.^26,27^ Comorbidities were considered present if the appropriate International Classification of Diseases, 9^th^ and 10^th^ Revisions codes (ICD-9 and ICD-10) were identified in the patient’s electronic health record. Comorbidities assessed in this analysis included: obesity, hypertension, prior myocardial infarction, prior bleeding, prior coronary artery bypass graft (CABG), congestive heart failure, coronary artery disease, diabetes mellitus, obstructive sleep apnea, stroke/transient ischemic attack, dyslipidemia, peripheral vascular disease, valvular heart disease, chronic kidney disease, obstructive pulmonary disease, alcohol abuse, drug use, dementia, and hyperthyroidism. Medications and vitals data within one year before or after the intervention were abstracted.

### Primary Outcomes

Our primary outcomes were prescription of anticoagulation and antiplatelet therapy. Prescription of anticoagulation therapy was evaluated according to stroke risk using the CHA_2_DS_2_-VASc score (High risk: ≥2 men or ≥3 women, Moderate risk: =1 men or =2 women, Low risk: =0 men or =1 women) and overall appropriateness of therapy defined as prescription of anticoagulation in high and moderate risk patients and no prescription of anticoagulation in low-risk patients. Prescription of antiplatelet therapy was evaluated according to the presence or absence of vascular disease and overall appropriateness defined as prescription of antiplatelet therapy in presence of vascular disease and no prescription of antiplatelet therapy in absence of vascular disease.

Each patient’s demographics and comorbidities were used to calculate their CHA_2_DS_2_-VASc score and medications data were used to determine if each patient was prescribed oral anticoagulation. Patients were considered as being prescribed oral anticoagulation (warfarin, direct thrombin inhibitor, direct factor Xa inhibitor) if they were started on an anticoagulant, were already prescribed an anticoagulant and did not discontinue the medication, or were restarted on an anticoagulant when they were seen by their primary care provider. Patients were not considered as being prescribed oral anticoagulation if they were not prescribed an anticoagulant or were discontinued from their anticoagulation therapy when they were seen by their primary care provider.

Vascular disease was defined as a known history of coronary artery disease, peripheral vascular disease, prior CABG, or prior myocardial infarction. Similar criteria as described above were used to determine if patients were prescribed, discontinued, or maintained on antiplatelet therapy (aspirin and/or P2Y12 inhibitor) during the time period that they were seen by their primary care provider.

### Secondary Outcomes

Secondary outcomes included the change in knowledge and self-reported confidence in AF management of providers participating in the intervention. Participants completed internally developed assessments of their knowledge of best practice recommendations for AF care and their confidence in treating AF, developed by the multi-disciplinary team of content experts based on key AF management recommendations from consensus guidelines.^23,24^ The knowledge survey was a case-based, 25 question assessment including questions on comorbidity management, stroke prophylaxis, and decisions on rate and rhythm control (Supplement). Scores ranged from 0-125, with higher scores indicating a higher knowledge base. The confidence survey assessed self-perceived ability to perform key objectives of the curriculum: EKG interpretation, assessment of the AF patient, patient education, comorbidity management, decisions on stroke prophylaxis, and AF medication management (Supplement). The assessment included 11 items based on the objectives of the 6-month intervention with confidence graded on a 5-point Likert scale. Scores ranged from 11-55 with higher scores indicating greater confidence in AF management. Participants completed both knowledge and confidence assessments before and after participation in the intervention.

### Statistical Analysis

Descriptive statistics were used to summarize frequency distributions for categorical variables and means, standard deviations for continuous variables in describing participant and patient characteristics, participant performance on the pre-intervention knowledge and confidence assessments, and rates of anticoagulation and antiplatelet therapy among patients. Changes in knowledge and confidence assessments before and after the intervention were assessed both overall and stratified by degree (MD, NP, PA) and specialty (cardiology, family medicine, internal medicine) using a Student’s t-test or one-way analysis of variance (ANOVA). Bivariable and multivariable linear regression were used to determine whether participant characteristics were associated with improvement in knowledge and confidence. Analyses of improvements in AF knowledge and confidence were limited to the participants who completed both the pre- and post-intervention assessments. Multivariable hierarchical logistic regression modeling was used to determine participant and patient predictors of oral anticoagulant use and antiplatelet use. Variables included in the multivariable model were identified from bivariable analyses. Statistical tests were 2-tailed, with p <0.05 considered significant. All statistical analyses were completed using R (2020, R Core Team, Vienna, Austria).

## RESULTS

Forty-one providers were recruited for participation in the intervention (Table 1). Of the 41 participants (64% physician, 29% nurse practitioner, 7% physician assistant), 49% worked in family medicine, 41% in internal medicine, and 10% in cardiology clinics. On average, participants were 16 years past completing their terminal degree. Participants attended an average of 14 (range: 4-22) educational sessions. On baseline questionnaires, the mean scores were 80 out of 125 maximum possible points in AF knowledge, and 28 out of 55 maximum possible points in AF confidence. Cardiology providers initially scored higher than family medicine and internal medicine providers (p=.003) on baseline AF knowledge. Baseline confidence in AF care was significantly lower among family medicine providers (p=.049).

**Table 1.**
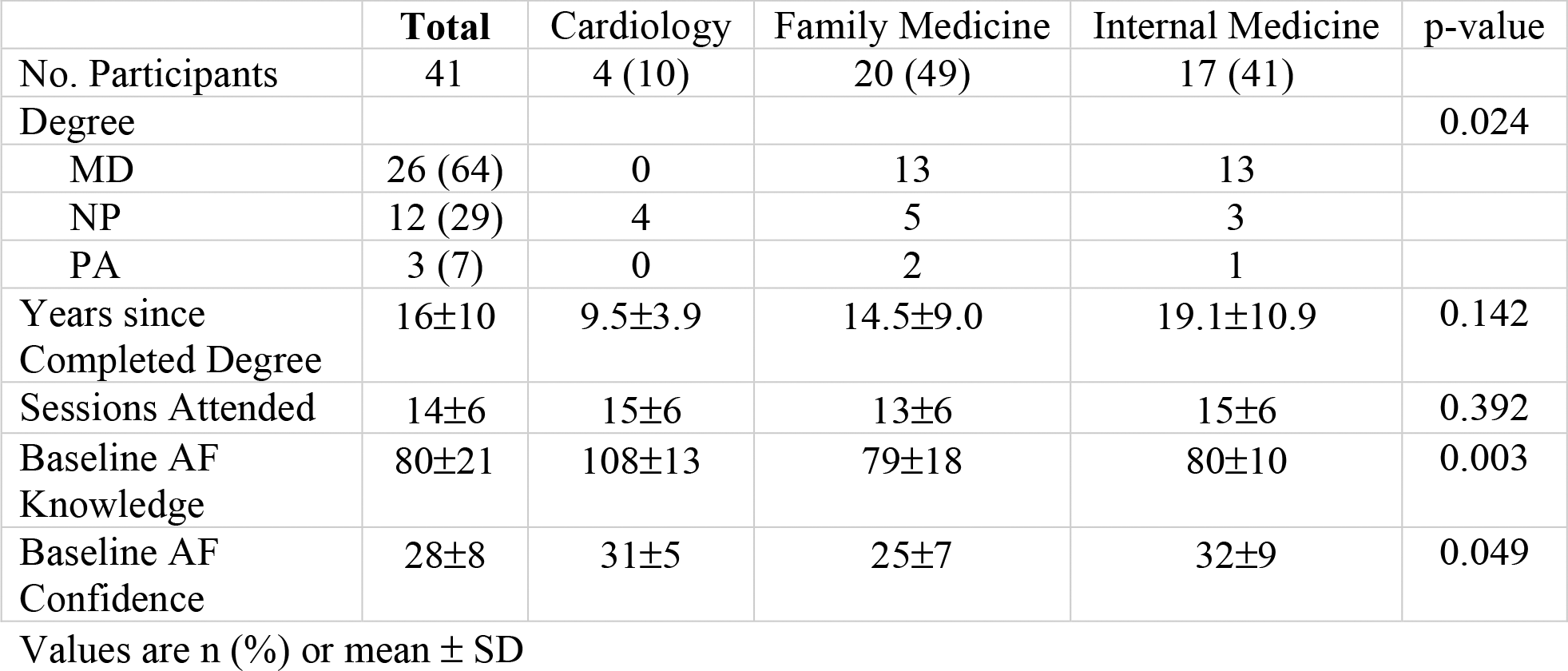
Baseline Characteristics of Participants.

|  | **Total** | **Cardiology** | **Family Medicine** | **Internal Medicine** | **p-value** |
| --- | --- | --- | --- | --- | --- |
| No. Participants | 41 | 4 (10) | 20 (49) | 17 (41) |  |
| Degree |  |  |  |  | 0.024 |
| MD | 26 (64) | 0 | 13 | 13 |  |
| NP | 12 (29) | 4 | 5 | 3 |  |
| PA | 3 (7) | 0 | 2 | 1 |  |
| Years since Completed Degree | 16±10 | 9.5±3.9 | 14.5±9.0 | 19.1±10.9 | 0.142 |
| Sessions Attended | 14±6 | 15±6 | 13±6 | 15±6 | 0.392 |
| Baseline AF Knowledge | 80±21 | 108±13 | 79±18 | 80±10 | 0.003 |
| Baseline AF Confidence | 28±8 | 31±5 | 25±7 | 32±9 | 0.049 |

Values are n (%) or mean ± SD

Baseline knowledge and confidence in AF care were similar across academic degree. During the pre-intervention period, participants saw 1,739 patients with AF and during the post-intervention period, participants saw 610 patients (Table 2). Compared to the post-intervention period, patients in the pre-intervention period were significantly older (p<.001) and more frequently self-identified their race as white (p<.001). Gender and SDI were not significantly different between patients in the two time periods. Patients in the post-intervention period had significantly higher rates of hypertension and diabetes (p<.001). There were no significant differences in CHA_2_DS_2_-VASc or ATRIA score, history of stroke or transient ischemic attack, bleeding, or vascular disease (CAD, CABG, MI, peripheral vascular disease) between the two groups.

**Table 2.** Patient Characteristics.

| Characteristic | Overall, N = 2,349<sup>1</sup> | Pre, N = 1,739<sup>1</sup> | Post, N = 610<sup>1</sup> | p-value<sup>2</sup> |
| --- | --- | --- | --- | --- |
| **Age** | 74.8 (11.31) | 75.3 (11.08) | 73.5 (11.85) | **<0.001** |
| **Gender** |  |  |  | 0.4 |
| Female | 1,138 (48%) | 834 (48%) | 304 (50%) |  |
| Male | 1,211 (52%) | 905 (52%) | 306 (50%) |  |
| **Race** |  |  |  | **<0.001** |
| Asian | 19 (0.8%) | 15 (0.9%) | 4 (0.7%) |  |
| Black | 367 (16%) | 247 (14%) | 120 (20%) |  |
| Other | 76 (3.2%) | 41 (2.4%) | 35 (5.7%) |  |
| White | 1,887 (80%) | 1,436 (83%) | 451 (74%) |  |
| **SDI** | 96.9 (21.00) | 96.6 (20.68) | 97.7 (21.88) | 0.7 |
| **BMI** | 30.1 (8.59) | 30.0 (8.70) | 30.4 (8.28) | 0.6 |
| Unknown | 458 | 353 | 105 |  |
| **Alcohol Abuse** | 26 (1.1%) | 22 (1.3%) | 4 (0.7%) | 0.2 |
| **h/o Bleeding** | 449 (19%) | 332 (19%) | 117 (19%) | >0.9 |
| **h/o CABG** | 256 (11%) | 200 (12%) | 56 (9.2%) | 0.11 |
| **h/o CAD** | 710 (30%) | 519 (30%) | 191 (31%) | 0.5 |
| **Congestive heart failure** | 40 (1.7%) | 30 (1.7%) | 10 (1.6%) | 0.9 |
| **COPD** | 605 (26%) | 449 (26%) | 156 (26%) | >0.9 |
| **TIA** | 321 (14%) | 230 (13%) | 91 (15%) | 0.3 |
| **Ischemic stroke** | 173 (7.4%) | 129 (7.4%) | 44 (7.2%) | 0.9 |
| **Hemorrhagic stroke** | 153 (6.5%) | 111 (6.4%) | 42 (6.9%) | 0.7 |
| **Dementia** | 76 (3.2%) | 55 (3.2%) | 21 (3.4%) | 0.7 |
| **Diabetes** | 399 (17%) | 249 (14%) | 150 (25%) | **<0.001** |
| **Drug use** | 65 (2.8%) | 45 (2.6%) | 20 (3.3%) | 0.4 |
| **Dyslipidemia** | 1,411 (60%) | 1,039 (60%) | 372 (61%) | 0.6 |
| **Hypertension** | 1,460 (62%) | 1,039 (60%) | 421 (69%) | **<0.001** |
| **Hyperthyroidism** | 66 (2.8%) | 47 (2.7%) | 19 (3.1%) | 0.6 |
| **h/o MI** | 339 (14%) | 245 (14%) | 94 (15%) | 0.4 |
| **Valvular heart disease** | 640 (27%) | 472 (27%) | 168 (28%) | 0.8 |
| **Peripheral vascular disease** | 548 (23%) | 395 (23%) | 153 (25%) | 0.2 |
| **CKD** | 319 (14%) | 229 (13%) | 90 (15%) | 0.3 |
| **Sleep Apnea** | 463 (20%) | 328 (19%) | 135 (22%) | 0.081 |
| **CHA2DS2_VASc Score** | 3.6 (1.74) | 3.6 (1.69) | 3.7 (1.89) | 0.3 |
| **ATRIA Score** | 2.7 (2.02) | 2.8 (2.03) | 2.6 (1.98) | 0.087 |

<sup>1</sup>Mean (SD); n (%)<sup>2</sup>Wilcoxon rank sum test; Pearson's Chi-squared test; Fisher's exact test

Rates of anticoagulant therapy stratified by CHA_2_DS_2_-VASc score, comparing the pre-intervention and post-intervention period are shown in Table 3. The majority (85%) of patients seen by participants during both time periods had high stroke risk (CHA_2_DS_2_-VASc score ≥2 men and ≥3 women), and the rate of anticoagulant use in the high-risk group following the training intervention increased from 35% to 46% (p<.001) (Figure 1). Among patients defined as low risk, there was also a significant increase in the rate of anticoagulant use from 19% to 37% (p=.033). The rate of therapy among patients with moderate CHA_2_DS_2_-VASc risk was unchanged following the training intervention. Overall, the rate of appropriate anticoagulant therapy increased from 37% to 46% (p<.001). In hierarchical multivariable logistic regression analyses (Table 5), the training intervention was significantly associated with appropriate use of anticoagulation overall (OR 1.4, p=.002) and in high-risk patients (OR 1.8, p<.001). In high-risk patients, older age was inversely associated with appropriate use of anticoagulation (OR 0.76, p<.001 per 10 yrs). Higher post-test score of knowledge was associated with appropriate use of anticoagulation overall and in high-risk patients, but not in low-risk patients. In low-risk patients, increased age and risk of bleeding (ATRIA score) were associated with appropriate anticoagulant use.

**Table 3.** Primary Outcome #1 – Anticoagulant Therapy Total Cohort.

| CHA<sub>2</sub>DS<sub>2</sub>-VASc | All |  | Pre |  | Post |  | p |
| --- | --- | --- | --- | --- | --- | --- | --- |
|  | N | Rate | n | Rate | n | Rate |  |
| High Risk | 1997 | 0.379 | 1489 | 0.351 | 508 | 0.461 | <0.001 |
| Moderate Risk | 234 | 0.338 | 170 | 0.318 | 64 | 0.391 | 0.293 |
| Low Risk | 118 | 0.246 | 80 | 0.188 | 38 | 0.368 | 0.033 |
| Overall Appropriate* | 2349 | 0.394 | 1739 | 0.369 | 610 | 0.464 | <0.001 |

\*Anticoagulant therapy was considered appropriate if prescribed in patients at high risk and moderate risk, or not prescribed in patients at low risk of stroke.

**Figure 1.**
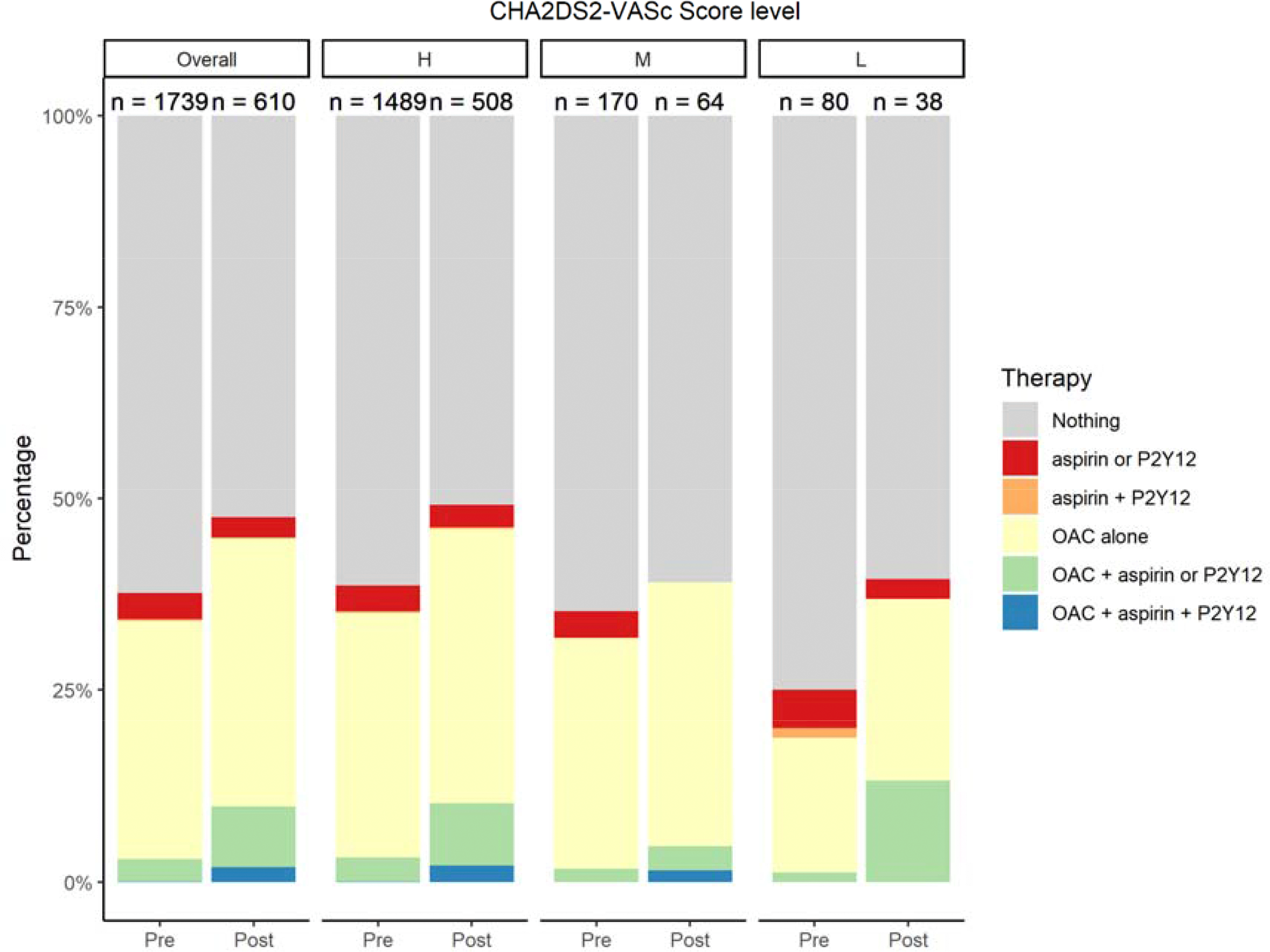
Medical therapy for stroke risk reduction of each CHA_2_DS_2_-VASc risk level and the overall patient cohort.

Absolute rates of antiplatelet therapy were low (Table 4), but increased significantly following the training intervention whether vascular disease was present or absent. Antiplatelet use in AF patients increased from 10% to 17% among patients with vascular disease and increased from 4% to 9% among patients without vascular disease. There were no differences in overall appropriateness of antiplatelet therapy before and after the intervention. Furthermore, in hierarchical multivariable logistic regression analyses (Table 6), the training intervention was not associated with change in use of antiplatelet therapy whether or not patients had vascular disease. Patients with vascular disease were less likely to receive appropriate antiplatelet therapy if they were older or female. Patients without vascular disease were less likely to receive antiplatelet therapy if they had hypertension or lower social deprivation index.

**Table 4.** Primary Outcome #2 - Antiplatelet Therapy Total Cohort.

| Vascular Disease | All |  | Pre |  | Post |  | p |
| --- | --- | --- | --- | --- | --- | --- | --- |
|  | N | Rate | n | Rate | n | Rate |  |
| Present | 1088 | 0.117 | 815 | 0.099 | 273 | 0.168 | 0.0021 |
| Absent | 1261 | 0.052 | 924 | 0.038 | 337 | 0.092 | <0.001 |
| Overall Appropriate* | 2349 | 0.563 | 1739 | 0.558 | 610 | 0.577 | <0.409 |

\*Antiplatelet therapy was considered appropriate if prescribed in patients with vascular disease and not prescribed in patients without vascular disease.

**Table 5.**
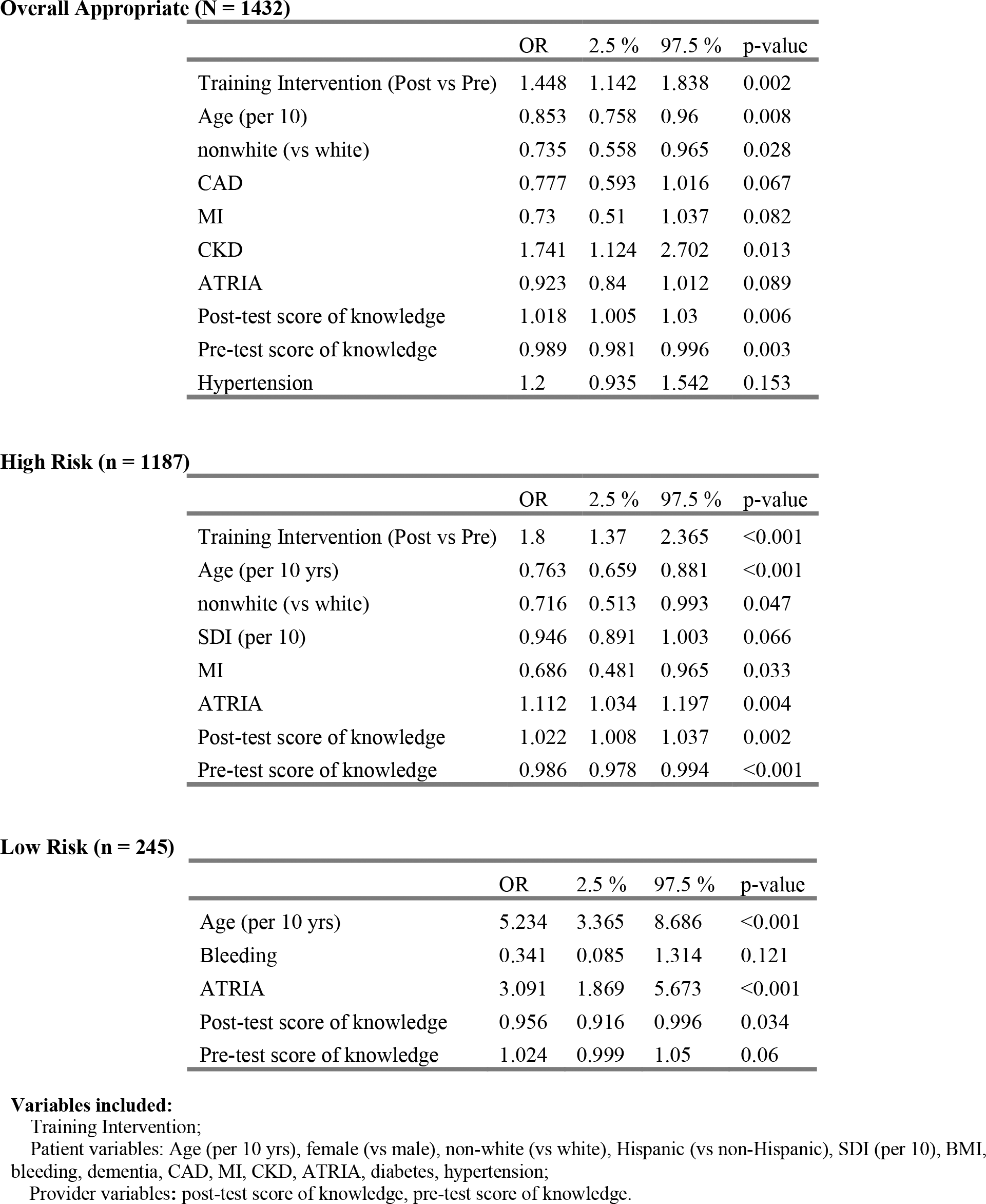
Hierarchical Model of Predictors and Covariates of Anticoagulation Therapy.

**Table 6.**
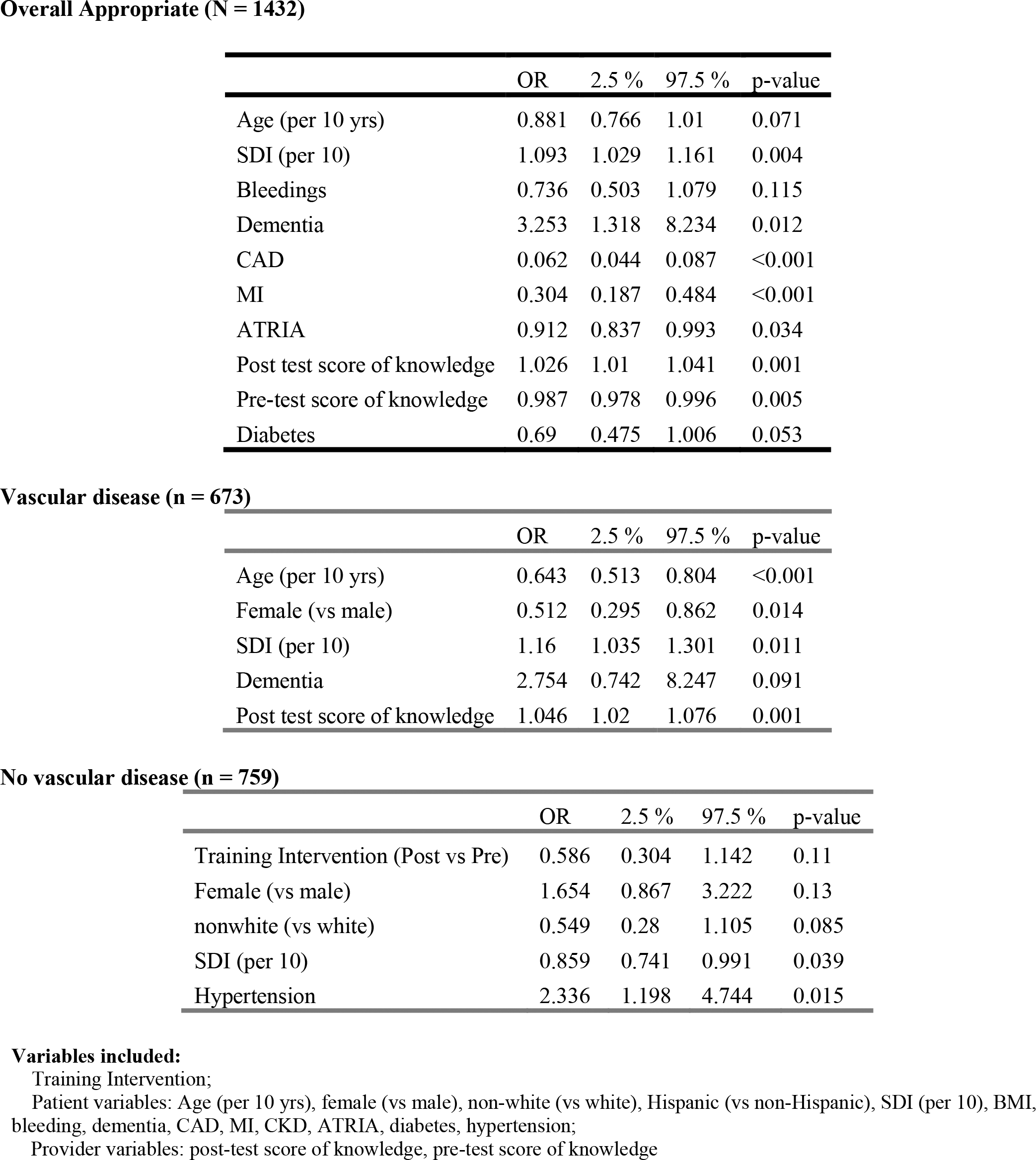
Hierarchical Model of Predictors and Covariates of Antiplatelet Therapy.

**Overall Appropriate (N = 1432)**
|  | OR | 2.5 % | 97.5 % | p-value |
| --- | --- | --- | --- | --- |
| Age (per 10 yrs) | 0.881 | 0.766 | 1.01 | 0.071 |
| SDI (per 10) | 1.093 | 1.029 | 1.161 | 0.004 |
| Bleedings | 0.736 | 0.503 | 1.079 | 0.115 |
| Dementia | 3.253 | 1.318 | 8.234 | 0.012 |
| CAD | 0.062 | 0.044 | 0.087 | <0.001 |
| MI | 0.304 | 0.187 | 0.484 | <0.001 |
| ATRIA | 0.912 | 0.837 | 0.993 | 0.034 |
| Post test score of knowledge | 1.026 | 1.01 | 1.041 | 0.001 |
| Pre-test score of knowledge | 0.987 | 0.978 | 0.996 | 0.005 |
| Diabetes | 0.69 | 0.475 | 1.006 | 0.053 |

**Vascular disease (n = 673)**
|  | OR | 2.5 % | 97.5 % | p-value |
| --- | --- | --- | --- | --- |
| Age (per 10 yrs) | 0.643 | 0.513 | 0.804 | <0.001 |
| Female (vs male) | 0.512 | 0.295 | 0.862 | 0.014 |
| SDI (per 10) | 1.16 | 1.035 | 1.301 | 0.011 |
| Dementia | 2.754 | 0.742 | 8.247 | 0.091 |
| Post test score of knowledge | 1.046 | 1.02 | 1.076 | 0.001 |

**No vascular disease (n = 759)**
|  | OR | 2.5 % | 97.5 % | p-value |
| --- | --- | --- | --- | --- |
| Training Intervention (Post vs Pre) | 0.586 | 0.304 | 1.142 | 0.11 |
| Female (vs male) | 1.654 | 0.867 | 3.222 | 0.13 |
| nonwhite (vs white) | 0.549 | 0.28 | 1.105 | 0.085 |
| SDI (per 10) | 0.859 | 0.741 | 0.991 | 0.039 |
| Hypertension | 2.336 | 1.198 | 4.744 | 0.015 |

The post-intervention questionnaires were completed by 28 (68%) of participants, and the individual improvement of each participant is shown in Figure 2. Overall, knowledge in AF care improved by 19 points (p<.001), and confidence in delivering AF care improved by 12 points (p<.001). Improvement in AF confidence was similar between specialties. In multivariable analysis, improvements in both knowledge (p=.021) and confidence (p=.002) were positively associated with the total number of sessions attended.

**Figure 2.**
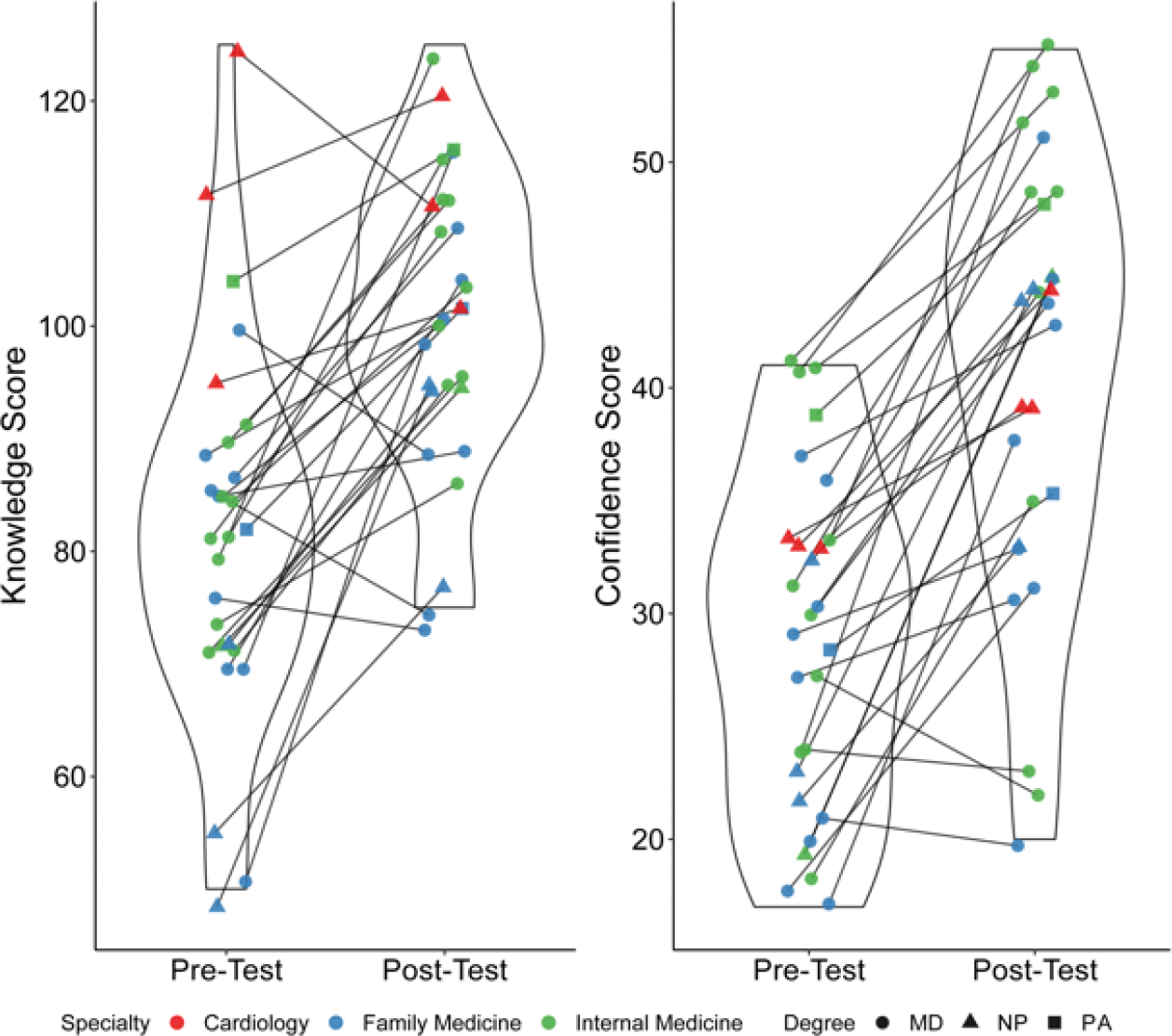
Individual improvement of each participant in AF knowledge and confidence on pre- and post-intervention questionnaires.

## DISCUSSION

This study evaluated the feasibility and benefit of a group-based virtual education intervention for primary care providers by measuring its impact on the use of stroke risk reduction strategies in AF patients and overall provider competency in AF care. Following a comprehensive 6-month intervention using the Project ECHO model, participating providers utilized more oral anticoagulation and antiplatelet therapy, particularly in patients at high risk of stroke. Participant knowledge and confidence in AF care also significantly improved.

Participation in the intervention was demonstrated to be the strongest driver of increased use of anticoagulation. Our study demonstrates the feasibility of a provider training intervention as a model to improve quality of care in communities where primary care providers may be the main resource for chronic cardiac disease management. Notably, we identified other demographic features which predict use of oral anticoagulation and antiplatelet therapy (age, race, gender). Understanding the drivers of these disparities of care will be important to help address this in future models of care.

Oral anticoagulation continues to be under-prescribed in AF patients with a high risk for stroke. A 2017 retrospective study found that among 94,000 patients with a history of AF who suffered from an ischemic stroke, 84% were not receiving therapeutic anticoagulation.^28^ Observational studies and systematic reviews demonstrate high rates of anticoagulant underuse, with over half of some cohorts being untreated for stroke risk reduction.^1,4,29,30^ A variety of factors are cited as being responsible for the lack of both prescription and adherence to anticoagulation, including socioeconomic status, comorbidities, and risk of bleeding of the patient, in addition to the priorities of the clinician who must provide recommendations that balance these factors.^31,32^ Common reasons that physicians may not prescribe anticoagulation are difficulty monitoring therapy and concerns about patient adherence.^5^ Our study confirms the presence of several patient demographic and clinical factors that may affect decisions on offering stroke prophylaxis. Current US guidelines recommend that anticoagulation be personalized based on shared decision making regarding the patient’s risk of stroke and bleeding, as well as life circumstances and preferences.^23^ Our study demonstrates that an ECHO model for educating primary care providers, who may be the sole provider caring for a majority of AF patients, on the role and options for therapy was effective in increasing rates of stroke risk reduction therapy and may be an effective option to elevate care in under-resourced communities in particular. However, there remains a need for further emphasis on evaluating risks and benefits of antithrombic therapy for specific patient populations such as elderly, non-white, and female patients or patients with comorbid cardiovascular disease and more education on the role of therapeutic options in patients with low risk for stroke and the role of antiplatelet therapy overall.

Our findings further demonstrate the feasibility and benefits of the ECHO model for spreading knowledge on complex arrhythmia management to community providers who may have more limited access to specialty care. Numerous Project ECHO studies have demonstrated the widespread benefits of training primary care providers to offer specialty care services, but few have applied this model to cardiac care and none specifically for AF. Management options for AF are rapidly evolving and the dissemination of best practices recommendations is challenging. After an initial study demonstrated that value of a Project ECHO model for treatment of hepatitis C, ^16^ many other ECHO studies have shown that provider knowledge and management skills can be improved across many diseases (e.g. hypertension, cancer, diabetes, dermatology, chronic pain management).^17–22^ Our study extends these observations and further supports translation of this educational model for treating other chronic diseases that require specialty care. Cardiology is a guideline-driven field with rapidly expanding technologies, and high-quality care depends on expert knowledge which may be more efficiently disseminated under the Project ECHO model. A project ECHO model can be a critical tool to alleviate disparities in care, particularly those driven by decreased access to specialty care.

Our results are also meaningful in the context of emerging models of care, including specialty AF clinics and AF centers of excellence. These initiatives have demonstrated improved quality of life, guideline adherence, and cost-effectiveness by reducing emergency department visits and hospitalizations for AF in addition to cardiovascular mortality.^11–14^ Similar to our study, specialty AF clinics have utilized a multidisciplinary approach, often including nurse-led care, to achieve these endpoints. However, the expenses associated with creating, staffing, and allocating physical resources to these clinics stands to be significantly reduced by using a virtual model. Furthermore, patients may still need to travel to these centers, which often exist in urban areas. Unlike these other options, our intervention equips primary care providers with the knowledge and confidence to address region-specific gaps and allows patients to receive high quality care within their own communities. Additionally, our model of care accommodates a wide range of AF providers and addresses the potential need for targeted, AF care pathways for rural populations. Limited provider knowledge and resources have been identified as contributors to poorer AF management in rural communities.^6^ Other studies have found that patients admitted to rural hospitals for AF had a 17% greater risk of death compared to urban centers, and elderly patients in rural areas are 12% less likely to receive antithrombotic therapy despite having significantly fewer relative contraindications.^7,9^ To our knowledge, no specific intervention has demonstrated the potential to bring AF care to under-resourced communities by elevating the level of care using embedded primary care providers.

### Limitations

There are several limitations with our study to consider. First, this is an observational study conducted at a single center that did not randomize participants to a control group. It is possible that participants would gradually develop the knowledge and confidence for AF management through other means. However, by staggering the cohorts of participants trained and the time frame utilized in the pre- and post-intervention comparison, there will be reduced time-dependent bias. Secondly, we observed an increase in the inappropriate use of oral anticoagulation therapy in patients with low stroke risk and inappropriate use of antiplatelet therapy in patients without vascular disease although these were in a small proportion of the patient population and not observed across all participating providers. Notably, there are potential indications for anticoagulation therapy in low-risk patients which were unmeasurable by our methodology (e.g., cardioversion, DVT prophylaxis, or recent catheter ablation). However, greater attention to education on the use of therapy in low-risk patients will be important to emphasize in future iterations of this intervention. Third, a challenge of this intervention was encouraging provider participation, including completion of the post-intervention knowledge and confidence surveys. Providers were incentivized to participate with continuing medical educational (CME) credits. Although providers participated in an average of 14 of 24 training sessions, there was a wide range with some providers participating in less than 10 sessions. Furthermore, only 28 out of 41 participants completed the post-intervention surveys despite numerous requests from the investigators. Thus, it is possible that those providers who benefitted less from the intervention were excluded from the secondary outcomes. However, of the 25 participants who attended the recommended 12 sessions, 22 (88%) completed the post-intervention surveys, indicating that our secondary outcomes are based on providers who completed the majority of the program. In future studies, it will be important to shorten the training intervention and consider additional incentive for participation. With greater participation, it is possible that the effect of the intervention may be enhanced. Lastly, the COVID-19 pandemic caused some disruptions to the intervention as some providers were reassigned to other duties. However, the virtual component allowed for continuation and completion of the intervention after a brief hiatus.

## CONCLUSIONS

Our findings suggest a virtual AF training intervention can significantly improve primary care provider competency in AF care and increase the adherence to best practice recommendations. Similar interventions may be useful to disseminate care for complex cardiac disease. Future studies can consider modifications to the training intervention and the potential for scaling this strategy to reach under-resourced communities in particular.

## Data Availability

According to the Transparency and Openness Promotion (TOP) Guidelines, access to data collected for this study will be made available upon reasonable request sent to the corresponding author. Because of the sensitive nature of the data, access will be considered for qualified researchers trained in human subject confidentiality protocols.

## ABBREVIATIONS

AF: Atrial fibrillation
CME: Continuing medical education
ECHO: Extension for community healthcare outcomes
EKG: Electrocardiogram
MD: Medical doctor
NP: Nurse practitioner
OAC: Oral anticoagulation
PA: Physician assistant
SDI: Social deprivation index

## FIGURE LEGENDS

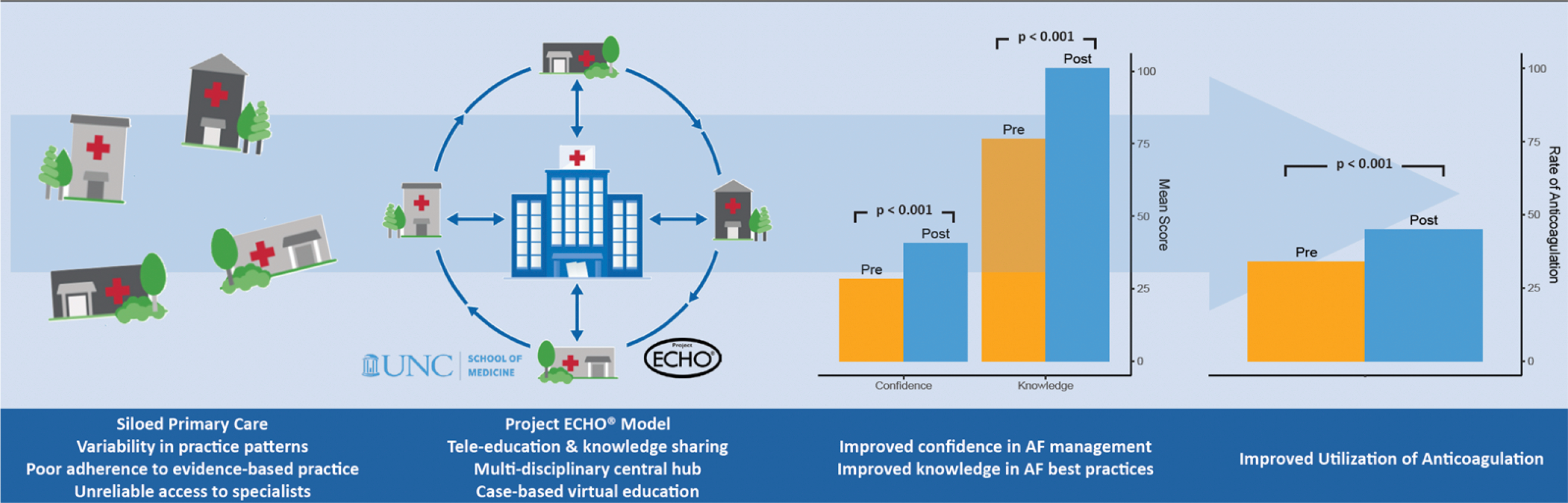

### Central Illustration

The Project ECHO model creates a “hub-and-spoke” knowledge-sharing network between specialists and regional care providers. Through case-based virtual education, primary care providers are able to become local AF experts and to provide high quality specialty care. At the provider level, knowledge and confidence in AF care significantly improved. There was also increased utilization of antithrombotic therapy among patients cared for by these providers after the training intervention.

